# Survey of National Practices for Postgraduate Year Two Critical Care and Emergency Medicine Pharmacy Residency Emergency Response Training

**DOI:** 10.1101/2023.01.22.23284886

**Authors:** Eli Philips, Clare Cycz, Ji T. Liu, I. Mary Eche, Adrian Wong

## Abstract

**Introduction:** Medical emergency response within the hospital involves an interdisciplinary team, including pharmacists. Pharmacist involvement in these teams has increased over time due to published benefits of their involvement. Development of emergency response skills may start during pharmacy residency, although limited data suggest how this is best implemented.

**Objectives:** Limited data evaluate post-graduate year 2 (PGY2) pharmacy resident training for emergency response, as well as PGY2 program values for this in screening/ranking candidates. A survey may help PGY2 programs improve their own programs and allow post-graduate year one (PGY1) pharmacy residents to identify residency programs that are a good fit for their career goals.

**Methods:** A list of PGY2 critical care (CC) and emergency medicine (EM) programs were identified. The questionnaire included program demographics, characteristics of PGY2 emergency response training, and PGY2 residency program values of PGY1 emergency response exposure for screening and ranking applicants for their programs. A Fisher’s exact test was used to compare differences between CC and EM programs for these outcomes.

**Results:** A total of 85 complete responses were analyzed (response rate: CC = 36.4%; EM = 32.1%). Emergency response training was often with both core and longitudinal experiences (72.9%), although differed by type of programs (p<0.001). Both CC and EM programs considered PGY1 pharmacy resident exposure to emergency response in screening candidates (33.9% and 57.7%, respectively), as well as ranking candidates (22% and 38.5%, respectively). For CC programs, both Advanced Cardiovascular Life Support certification and quantity of PGY1 emergency response ranked as the most important characteristics, while EM programs ranked quantity as the most important.

**Conclusion:** The results of this survey indicate heterogeneity in PGY2 CC and EM emergency response training. PGY1 applicants for these programs should consider their experience with emergency response as a factor in identifying an appropriate program for their initial training.

## INTRODUCTION

Medical emergency response within the hospital typically involves an interdisciplinary team of clinicians with different expertise, which has been suggested as an intervention by the Institute for Healthcare Improvement to improve patient outcomes. [1] Pharmacists can be valuable members of the medical response team given their expertise in medication therapy. [2] A United States hospital analysis in 1998 found that pharmacist participation in cardiopulmonary resuscitation (CPR) teams was associated with a reduction of 12880 deaths per year. [3] This reduction in mortality is likely due to a reduction in medication errors, which are common in these situations. [4-6] Pharmacist participation in these events has increased from 2012 to 2021, with an increase in rapid response team and code team participation increasing from 19.1% to 36% and 41.1% to 69.7%, respectively. [7]

Pharmacist training in emergency response is crucial to ensure medication recommendations, medication preparation, and provision of drug information are optimized. [8,9] Pharmacy residency facilitates the development of skills in emergency response, especially for learners who are interested in critical care (CC) or emergency medicine (EM), where these skills are mandatory. A survey of postgraduate year one (PGY1) and postgraduate year two (PGY2) pharmacy residency training programs published in 2007 found that 30% of respondents had required pharmacy resident participation in CPR teams, with 38% of respondents having this optional for residents. [10] This survey found that many methods were used to improve resident skills in emergency response, including simulation and didactic lectures. Limited data exist evaluating how PGY2 pharmacy residents are trained to become independent emergency responders, which is a key objective of accreditation by pharmacy professional organizations. A key objective of CC and EM PGY2 pharmacy residents’ training programs is achieving independent medical emergency response. Despite this, there is a lack of data evaluating how PGY2 pharmacy residents are trained in this area.

Professional societies have supported pharmacist emergency response services with official statements and the American Society of Health-System Pharmacists (ASHP) has noted these skills as required objectives during PGY2 residencies specializing in CC or EM. [11-12] However, limited data exist evaluating what PGY2 residency programs value in prospective residents in regards to emergency response training. This information may help PGY1 pharmacy residents identify residency programs that align with their training and career goals. A survey of PGY2 residency programs was developed to determine current practices and areas for improvement for emergency response training.

## METHODS

### Survey Development and Administration

An anonymous, electronic survey was developed to describe characteristics of PGY2 pharmacy resident emergency response, as well as PGY2 residency program values when evaluating PGY1 pharmacy resident exposure to emergency response. A contact list of PGY2 residency program directors (RPDs) for CC and EM programs was developed using information from the ASHP and American College of Clinical Pharmacy residency directories, with contact information verified with the corresponding residency website. The electronic Qualtrics survey was sent in May 2022, with a final deadline of June 2022. One response was requested from a representative of each program. Survey requests that were not deliverable or incomplete responses were not included in our analysis. This survey was approved by the Beth Israel Deaconess Medical Center Institutional Review Board.

The questionnaire (see Supplementary Information A) included demographic information, characteristics of PGY2 pharmacy resident emergency response, including training, extent of emergency response exposure, and determination of ability to independently respond to emergencies (as rated by the representative), and the importance of PGY1 emergency response exposure to the PGY2 residency program in terms of screening and ranking candidates. A 5-point Likert scale (1 = most important; 5 = least important) was used to characterize the importance of PGY1 pharmacy resident training in terms of PGY2 residency program values for screening/ranking candidates. A free-text response was available to determine beneficial resources at their program in developing their pharmacy resident emergency response program.

### Statistical analysis

Survey responses were analyzed using descriptive statistics. A Fisher’s exact test was used to compare PGY2 residency program values in PGY1 pharmacy resident emergency response exposure and characteristics of PGY2 pharmacy resident emergency response between CC and EM programs. Statistical analyses were performed using R (version 4.2.1; Vienna, Austria), with a two-sided p-value of less than 0.05 considered to indicate statistical significance.

## RESULTS

A total of 256 programs were contacted, with 243 meeting inclusion criteria (CC: n=162; EM: n=81). Of those included, there were 85 completed responses (response rate: CC 36.4%; EM 32.1%). Programs included in the analysis were predominately at an academic medical center, with at least 250 beds (Table 1).

**Table 1.** Respondent Institution Characteristics

| **Table 1. Respondent Institution Characteristics** |  |  |
| --- | --- | --- |
|  | **CC (n=59)** | **EM (n=26)** |
| Type of Institution, n (%) |  |  |
| Government | 2 (3.4) | 0 (0) |
| Community hospital, Non-teaching | 4 (6.8) | 6 (23.1) |
| Community hospital, Teaching | 16 (27.1) | 6 (23.1) |
| Academic medical center | 37 (62.7) | 14 (53.8) |
| Number of Hospital Beds, n (%) |  |  |
| <50 | 0 (0) | 0 (0) |
| 50-149 | 1 (1.7) | 0 (0) |
| 150-249 | 0 (0) | 0 (0) |
| 250-499 | 15 (25.4) | 7 (26.9) |
| 500-750 | 21 (35.6) | 9 (34.6) |
| >750 | 22 (37.3) | 10 (38.5) |
| Number of ED Beds, n (%) |  |  |
| None | 1 (1.7) | 0 (0) |
| 1-19 | 0 (0) | 0 (0) |
| 20-49 | 7 (11.9) | 9 (34.6) |
| 50-74 | 24 (40.7) | 9 (34.6) |
| 75-100 | 16 (27.1) | 5 (19.2) |
| >100 | 11 (18.6) | 3 (11.5) |
| Number of ICU Beds, n (%) |  |  |
| 0-12 | 0 (0) | 0 (0) |
| 13-25 | 1 (1.7) | 2 (7.7) |
| 26-49 | 7 (11.9) | 7 (26.9) |
| 50-74 | 11 (18.6) | 2 (7.7) |
| 75-100 | 13 (22.0) | 5 (19.2) |
| >100 | 27 (45.8) | 10 (38.5) |
| Age of Residency Program, n (%) |  |  |
| <1 year | 1 (1.7) | 2 (7.7) |
| 1-3 years | 5 (8.5) | 3 (11.5) |
| 4-7 years | 15 (25.4) | 14 (53.8) |
| 8-10 years | 7 (11.9) | 4 (15.4) |
| >10 years | 31 (52.5) | 3 (11.5) |
| Pharmacist Emergency Response, n (%) |  |  |
| Sepsis | 14 (23.7) | 9 (34.6) |
| Pediatric | 29 (49.2) | 21 (80.8) |
| Intubation/Airway | 35 (59.3) | 21 (80.8) |
| Trauma | 43 (72.9) | 22 (84.6) |
| Stroke | 53 (89.8) | 26 (100) |
| Code blue | 58 (98.3) | 26 (100) |
| Other | 2 (3.4) | 6 (23.1) |
| PGY2 Resident Emergency Response, n (%) |  |  |
| Sepsis | 10 (16.9) | 9 (34.6) |
| Pediatric | 23 (39.0) | 21 (80.8) |
| Intubation/Airway | 32 (54.2) | 23 (88.5) |
| Trauma | 37 (62.7) | 22 (84.6) |
| Stroke | 47 (79.7) | 26 (100) |
| Code blue | 58 (98.3) | 26 (100) |
| Other | 3 (5.1) | 6 (23.1) |

CC = critical care; ED = emergency department; EM = emergency medicine; ICU = intensive care unit; PGY2 = postgraduate year two

Critical care programs were generally more established, with >50% in existence for over 10 years, while EM programs were typically established for 4-7 years. For all respondents, both pharmacists and PGY2 pharmacy residents participate in emergency response at their institution, predominately in code blue and stroke. The ‘other’ responses, which were infrequent, included code violet (i.e., violent patient) (n=2), malignant hyperthermia, massive transfusion protocol, obstetrics, procedural sedation (n=2), pulmonary embolism response team, ST-elevation myocardial infarction activations (n=3), therapeutic hypothermia response, and toxicologic emergencies (see Supplementary Information B).

### Characteristics of PGY2 Pharmacy Resident Training

The majority of both CC and EM programs did not have an on-call program (CC: 59.3%; EM: 73.1%). Emergency response for PGY2s was often both a core and longitudinal experience during their residency year, which was different between CC and EM programs (Table 2).

**Table 2.** Characteristics of PGY2 Emergency Response

| Table 2. Characteristics of PGY2 Emergency Response |  |  |  |
| --- | --- | --- | --- |
|  | CC (n=59) | EM (n=26) | p-value |
| PGY2 Emergency Response Experience, n (%) |  |  |  |
| Core experience only | 13 (22.0) | 7 (26.9) | <0.001 |
| Longitudinal experience only | 2 (3.4) | 1 (3.8) |  |
| Both core and longitudinal experience | 44 (74.6) | 18 (69.2) |  |
| Types of Training, n (%) |  |  |  |
| PALS | 25 (42.4) | 20 (76.9) | N/A |
| Interdisciplinary simulation | 30 (50.8) | 17 (65.4) |  |
| Mannequin simulation | 33 (55.9) | 13 (50.0) |  |
| Pairing w/ preceptor | 43 (72.9) | 20 (76.9) |  |
| ACLS | 55 (93.2) | 25 (96.2) |  |
| Hands-on training | 57 (96.6) | 26 (100) |  |
| Other | 3 (5.1) | 5 (19.2) |  |
| Independent Response Determination, n (%) |  |  |  |
| Resident do not independently respond | 6 (10.2) | 1 (3.8) | N/A |
| Set time during residency year | 8 (13.6) | 4 (15.4) |  |
| Meeting threshold of responses | 14 (23.7) | 5 (19.2) |  |
| Passing grade on formal evaluation | 15 (25.4) | 2 (7.7) |  |
| Preceptor sign-off | 38 (64.4) | 25 (96.2) |  |
| Other | 12 (20.3) | 2 (7.7) |  |
| Independent Response Timeline, n (%) |  |  |  |
| Beginning of residency-August | 13 (22.0) | 3 (11.5) | 0.584 |
| September-October | 18 (30.5) | 11 (42.3) |  |
| November-December | 11 (18.6) | 9 (34.6) |  |
| January-February | 10 (16.9) | 3 (11.5) |  |
| March-April | 1 (1.7) | 0 (0) |  |
| May-End of residency | 1 (1.7) | 0 (0) |  |
| Never able to respond independently | 3 (5.1) | 0 (0) |  |
| Number of PGY2 Emergency Responses by End of Residency Year, n (%) |  |  |  |
| 1-25 | 10 (16.9) | 0 (0) | <0.001 |
| 26-50 | 23 (39.0) | 2 (7.7) |  |
| 51-99 | 17 (28.8) | 6 (23.1) |  |
| 100-149 | 5 (8.5) | 6 (23.1) |  |
| 150-199 | 4 (6.8) | 3 (11.5) |  |
| >200 | 0 (0) | 9 (34.6) |  |

ACLS = Advanced Cardiovascular Life Support; CC = critical care; EM = emergency medicine; PALS = Pediatric Advanced Life Support; PGY2 = postgraduate year two

The type of training was variable, with ‘other’ responses including Advanced Trauma Life Support (n=3), Emergency Neurological Life Support (n=2), Pediatric Fundamental Critical Care Support, and Neonatal Resuscitation Program certification. Preceptor sign-off was the predominant method of determining a PGY2 pharmacy resident’s ability to respond independently, with the majority of residents being able to do so by October of their residency year. The quantity of emergency responses reported that PGY2 pharmacy residents had attended by the end of their residency year was significantly higher in EM programs than CC programs (p<0.001)

### PGY2 Residency Program Values for PGY1 Pharmacy Resident Emergency Response

PGY2 residency programs placed different emphasis on the importance of PGY1 emergency response exposure, with 30% of CC and 60% of EM programs factoring this into screening PGY2 applicants (p=0.056) (Table 3).

**Table 3.** Programs Identifying Importance of PGY1 Emergency Response in Screening or Ranking Candidates for PGY2 Program

|  |  |  |  |
| --- | --- | --- | --- |
|  | **CC (n=59)** | **EM (n=26)** | **p-value** |
| PGY1 Exposure for Screening, n (%) | 20 (33.9) | 15 (57.7) | 0.056 |
| Weight of <5% | 6 (30.0) | 3 (11.5) | 0.048 |
| Weight of 5-10% | 10 (50.0) | 2 (3.4) |  |
| Weight of 10-15% | 2 (10.0) | 5 (3.4) |  |
| Weight of 15-20% | 2 (10.0) | 3 (11.5) |  |
| Weight of >20% | 0 (0) | 2 (7.7) |  |
| PGY1 Exposure for Ranking, n (%) | 13 (22.0) | 10 (38.5) | 0.184 |

CC = critical care; EM = emergency medicine; PGY1 = postgraduate year one

The weight of this was relatively low (<10%) for CC programs and variable for EM programs (p=0.048). This experience was more considered for screening than ranking candidates for both CC and EM programs. When ranking candidates, EM programs had a higher consideration for this experience than CC programs, although this was not statistically significant (p=0.184).

Several factors are reported to delineate how PGY2 programs evaluate PGY1 emergency response in screening/ranking candidates. For CC programs, Advanced Cardiovascular Life Support (ACLS) certification was the most considered characteristic, followed by participation in a PGY1 on-call program (Table 4).

**Table 4.**
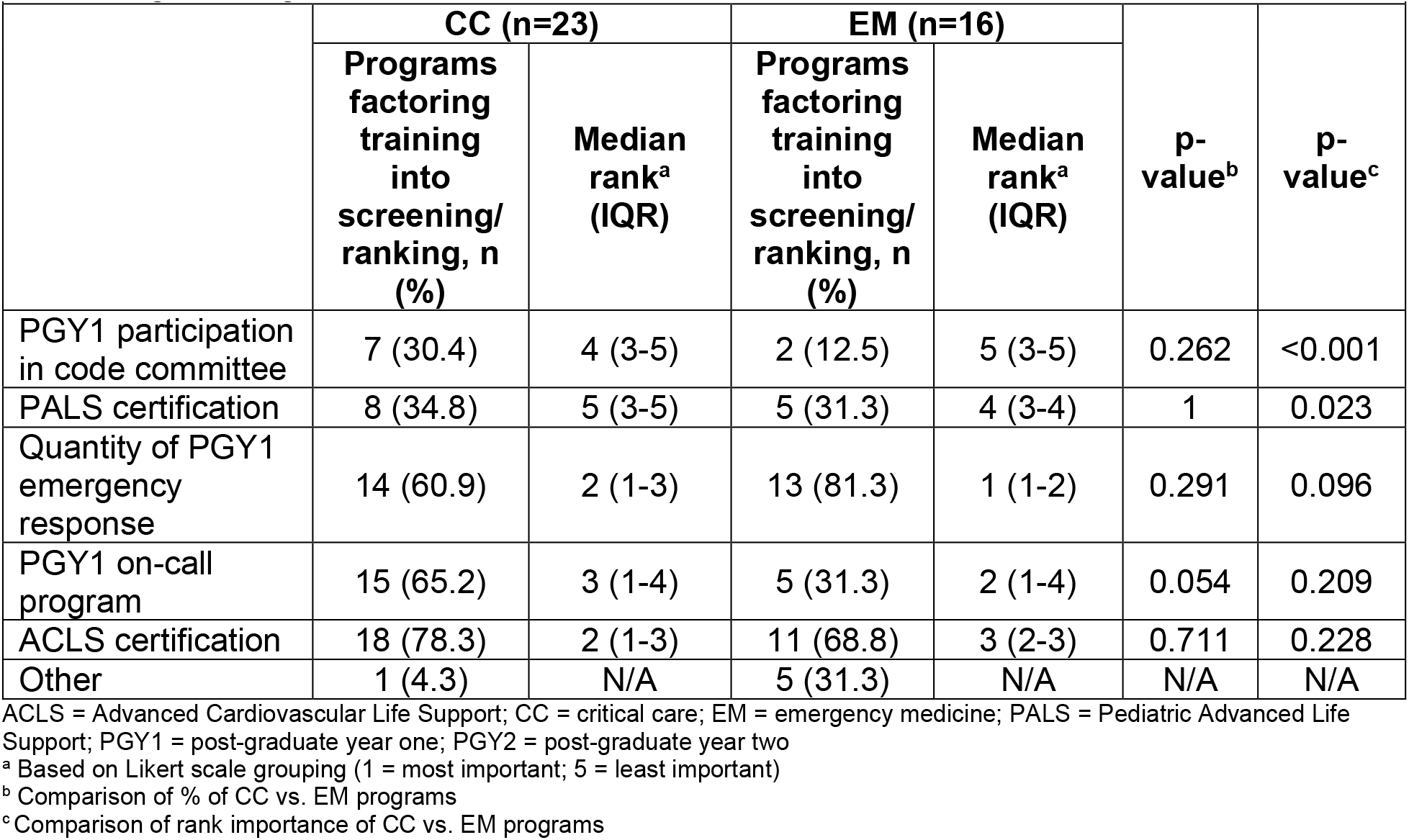
Characteristics of PGY1 Emergency Response for PGY2 Program Screening/Ranking

|  |  |  |  |  |  |  |
| --- | --- | --- | --- | --- | --- | --- |
|  | **CC (n=23)** |  | **EM (n=16)** |  | **p-value<sup>b</sup>** | **p-value<sup>c</sup>** |
|  | **Programs factoring training into screening/ranking, n (%)** | **Median rank<sup>a</sup> (IQR)** | **Programs factoring training into screening/ranking, n (%)** | **Median rank<sup>a</sup> (IQR)** |  |  |
| PGY1 participation in code committee | 7 (30.4) | 4 (3-5) | 2 (12.5) | 5 (3-5) | 0.262 | <0.001 |
| PALS certification | 8 (34.8) | 5 (3-5) | 5 (31.3) | 4 (3-4) | 1 | 0.023 |
| Quantity of PGY1 emergency response | 14 (60.9) | 2 (1-3) | 13 (81.3) | 1 (1-2) | 0.291 | 0.096 |
| PGY1 on-call program | 15 (65.2) | 3 (1-4) | 5 (31.3) | 2 (1-4) | 0.054 | 0.209 |
| ACLS certification | 18 (78.3) | 2 (1-3) | 11 (68.8) | 3 (2-3) | 0.711 | 0.228 |
| Other | 1 (4.3) | N/A | 5 (31.3) | N/A | N/A | N/A |

ACLS = Advanced Cardiovascular Life Support; CC = critical care; EM = emergency medicine; PALS = Pediatric Advanced Life Support; PGY1 = post-graduate year one; PGY2 = post-graduate year two
<sup>a</sup> Based on Likert scale grouping (1 = most important; 5 = least important)
<sup>b</sup> Comparison of % of CC vs. EM programs
<sup>c</sup> Comparison of rank importance of CC vs. EM programs

For EM programs, the quantity of PGY1 pharmacy resident emergency response was the most considered characteristic, followed by ACLS certification. There were no statistically significant differences between these characteristics between CC and EM programs. In terms of rank order of importance, both ACLS certification and quantity of PGY1 pharmacy resident emergency response were ranked highest, while quantity was the most important characteristic for EM programs. When comparing this ranking between CC and EM programs, PGY1 pharmacy resident participation in a code committee was ranked higher IN CC programs than EM programs (p<0.001), while Pediatric Advanced Life Support was ranked higher by EM programs (p=0.023).

## DISCUSSION

In our survey evaluating PGY2 CC and EM residency programs, we found that residents of these programs consistently engage in emergency response, although there are differences between CC and EM programs in terms of extent of emergency response experience and emphasis of PGY1 pharmacy emergency response when screening and ranking candidates. Our data suggest that more objective guidance of emergency response competency from professional organizations may be needed given the observed heterogeneity in terms of emergency response training of PGY2 CC and EM residents.

The ASHP Practice Advancement Initiative 2030 recommends that pharmacists participate in key roles in emergency response. [13] Examples of benefits that pharmacists have shown in emergency response include a reduction in time to fibrinolytic in stroke, reduction in time to antimicrobials for patients with suspected sepsis, and a decrease in medication errors during cardiopulmonary resuscitation. [14-16] It is important that CC and EM pharmacy residents develop these skills to ensure medication recommendations, medication preparation, and provision of drug information are optimized. [17-19]

The training of PGY2 residents has shown progression since the findings of a survey published in 2007. [10] This survey found that 21% of respondents provided core rotation-specific training for emergency response, while only three programs provided longitudinal development. We found that emergency response was emphasized in both core and longitudinal experiences in the majority of programs. Longitudinal development is important, as a study that evaluated the integration of pharmacy residents within an interdisciplinary simulation has been shown to improve skills, but these skills may degrade over a period of one month if not reinforced. [20] Potential areas of focus, outside clinical knowledge for developing skills may be based on certain themes, such as communication, adaptability, analytics, focus, and responsibility. [21] It was worth noting that some programs reported that PGY2 pharmacy residents are not able to respond independently by the end of the residency. This indicates the need for further studies to identify best practices for developing emergency response skills.

The 2007 survey also found that the most common evaluation method for determining ability to independently respond were mock simulations, followed by an objective evaluation via written test or direct observation of an emergency response. [10] This differed from our findings that preceptor sign-off was the most common method, a technique which may be subject to variability based on institution, preceptor, type of medical emergency and other factors. Future characterization of what is considered when determining if PGY2 pharmacy residents may respond independently would assist in standardization across programs. Additionally, use of simulation training was common, which has been shown to increase awareness of pharmacist roles, responsibilities, and emergency skills. [22,23] We found that all PGY2 EM residency programs were confident in the ability of their residents to respond independently by February of their residency year, while this was more variable for CC programs. Emergency medicine programs also had significantly more experience with emergency response than CC programs. Given emergency response is a required competency for ASHP accreditation, more objective guidance on training and determination of competency may be helpful for both CC and EM PGY2 programs. For programs that may not have certain resources for training, collaboration with other programs (e.g., regional) is a potential method to address this limitation.

The results of this survey found that both CC and EM PGY2 residency programs considered PGY1 emergency response in screening prospective candidates. The weight of these experiences was low in CC programs but was more variable in EM programs. Therefore, PGY1 emergency response experiences may be one of many considerations in determining an appropriate program for the candidate, especially if they are interested in pursuing a PGY2 in EM. It was interesting to see that quantity of emergency response was an important factor for both CC and EM programs. This seems difficult to ascertain from an application outside of rotations and longitudinal activities, with emergency response data unlikely to be provided in a quantitative form. After interviewing a candidate, this seems to be more likely to be evaluated while ranking candidates.

Our study had limitations, including the response rate, not including PGY2 pediatric residency programs, the potential for one institution to have responses from both CC and EM programs, and our focus on the perspective of the residency program and not the resident. Our response rate for both CC and EM programs was approximately 30%, which was similar to the survey published in 2007. [10] Although this may be considered a low response rate, we believe that the heterogeneity in responses supports that we had a representative sample of programs and practices. Our focus was on adult patients, given our study team’s expertise. Therefore, the lack of inclusion of PGY2 pediatrics residency programs limits our findings to this population, although a significant proportion of programs did have PALS certification, namely within EM programs. Although we attempted to restrict responses to one per CC or EM program, there likely are institutions that have both CC and EM programs, which would have provided similar demographic data. However, given the design of CC and EM programs often differs, we believe this would not affect the interpretation of our results. Finally, given the lack of standardized training expectations of PGY2 pharmacy resident emergency response our data is likely subject to recall bias and variation given subjectivity of resident evaluation. Our data only reflect the perspective of the residency program and inclusion of resident self-evaluation of performance and competency would add additional insight.

## CONCLUSIONS

The results of this survey indicate variability in developing PGY2 CC and EM pharmacy residents in terms of emergency response skills and may help certain programs identify potential areas of improvement. Our data suggest that PGY1 pharmacy residents interested in pursuing PGY2 CC or EM training should consider emergency response training as a factor in identifying PGY1 programs, especially if they are interested in EM. Future studies should help to identify best practices for developing and evaluating emergency response skills.

## Data Availability

All data produced in the present study are available upon reasonable request to the authors

## Acknowledgements

There was no funding, financial, or material support for this research

