## Supplementary Information for "Survey of National Practices for Postgraduate Year Two Critical Care and Emergency Medicine Pharmacy Residency Emergency Response Training"

SUPPLEMENTARY APPENDIX

**Appendix A. Questionnaire**

PGY2 Pharm Res Code Competency

Q2.1 Which of the following best reflects your role in the PGY2 residency program at your institution?

- Residency Program Coordinator (1)
- Residency Program Director (2)
- Preceptor (4)
- Other (specify) (3) __________________________________________________

Q2.2 Which of the following best reflects which type of PGY2 residency program you are representing?

- Critical Care (1)
- Emergency Medicine (2)

Q2.3 Which of the following best describes your institution?

- Academic medical center (1)
- Community hospital, teaching (2)
- Community hospital, non-teaching (3)
- Government (e.g., VA) (4)
- Other (specify) (5) __________________________________________________

Q2.4 How many hospital beds does your institution have?

- <50 beds (1)
- 50-149 beds (5)
- 150-249 beds (6)
- 250-499 beds (2)
- 500-750 beds (3)
- >750 beds (4)

Q2.5 How many licensed ED beds does your institution have?

- No ED (1)
- 1-19 beds (2)
- 20-49 beds (4)
- 50-74 beds (5)
- 75-100 beds (7)
- Greater than 100 beds (6)

Q2.6 How many ICU beds does your institution have?

- 0-12 beds (1)
- 13-25 beds (2)
- 26-49 beds (3)
- 50-74 beds (4)
- 75-100 beds (5)
- Greater than 100 beds (6)

Q2.7 How long has your PGY2 residency program been in existence for?

- Less than 1 year (1)
- 1-3 years (2)
- 4-7 years (3)
- 8-10 years (4)
- More than 10 years (6)

Q2.8 Which of the following best defines your ***PGY2 on-call program***?

- On-site, AM/PM (1)
- On-site, 24/7 (2)
- Out-of-hospital, AM/PM (4)
- Out-of-hospital, 24/7 (5)
- No on-call program (3)
- Other (specify) (6) __________________________________________________

| Page Break |
| --- |

Q2.9 Which of the following describes the type(s) of medical emergencies that ***pharmacists*** respond to at your institution? [**select all that apply**]

- Code Blue (1)
- Code Stroke (2)
- Code Sepsis (3)
- Code Trauma (4)
- Intubation/Airway (5)
- Pediatric emergency response (8)
- Rapid response team (6)
- Other (specify) (7) __________________________________________________

Q2.10 Pharmacy residents in my PGY2 program actively participate (e.g., medication prep, clinical recommendations) in medical emergency response (e.g., Code Blue, stroke).

- Yes (1)
- No (2)

Skip To: End of Block If Pharmacy residents in my PGY2 program actively participate (e.g., medication prep, clinical recom... = No

| Page Break |
| --- |

Q3.1 Is PGY1 resident exposure to medical emergency response currently one of your evaluation criteria considered in **screening candidates for interviews** for your PGY2 program?

- Yes (1)
- No (2)

Q3.2 Is PGY1 resident exposure to medical emergency response currently one of your evaluation criteria considered in **ranking candidates** for your PGY2 program?

- Yes (1)
- No (2)

| Page Break |
| --- |

Display This Question:

If Is PGY1 resident exposure to medical emergency response currently one of your evaluation criteria... = Yes

Or Is PGY1 resident exposure to medical emergency response currently one of your evaluation criteria... = Yes

Q3.3 Which of the following do you consider in identifying candidates (screening/ranking) for your PGY2 residency? [**select all that apply**]

- ACLS certification (1)
- PALS certification (2)
- PGY1 on-call program (3)
- PGY1 participation in code committee (4)
- Quantity of PGY1 emergency response experiences (5)
- None of the above (6)
- Other (specify) (7) __________________________________________________

Display This Question:

If Is PGY1 resident exposure to medical emergency response currently one of your evaluation criteria... = Yes

Or Is PGY1 resident exposure to medical emergency response currently one of your evaluation criteria... = Yes

Q3.4 Please rank the following emergency response experiences from most important (top row, #1) to least important (bottom row, #5) in identifying candidates for your PGY2 program.

______ ACLS certification (1)

______ PALS certification (2)

______ PGY1 on-call program (3)

______ PGY1 participation in code committee (4)

______ Quantity of PGY1 emergency response experiences (5)

| Page Break |
| --- |

Display This Question:

If Is PGY1 resident exposure to medical emergency response currently one of your evaluation criteria... = Yes

Q3.5 Which of the following best describes the weight that PGY1 emergency medical response experience has in screening candidates for interviews in your PGY2 program?

- Less than 5% (1)
- 5-10% (2)
- 10-15% (3)
- 15-20% (4)
- Greater than 20% (5)

| Page Break |
| --- |

Display This Question:

If Pharmacy residents in my PGY2 program actively participate (e.g., medication prep, clinical recom... = Yes

Q5.1 Which of the following describes the type(s) of medical emergencies that **pharmacy residents** respond to at your institution? [**select all that apply**]

- Code Blue (1)
- Code Stroke (2)
- Code Sepsis (3)
- Code Trauma (4)
- Intubation/Airway (5)
- Pediatric emergency response (8)
- Rapid response team (6)
- Other (specify) (7) __________________________________________________

Q5.2 PGY2 residents have experience with medical emergency response during the following experiences: [**select all that apply**]

- Longitudinal learning experience (1)
- Core learning experience (e.g., ICU, ED) (2)
- Other (specify) (3) __________________________________________________

| Page Break |
| --- |

Q5.3 Which of the following ***training*** do PGY2 pharmacy residents complete at your institution to ***prepare them for code response***? [**select all that apply**]

- ACLS certification (1)
- Hands-on training with code cart (2)
- Interdisciplinary simulation (outside of ACLS certification) (5)
- Mannequin simulation (3)
- Pairing resident with preceptor (4)
- PALS certification (8)
- None of the above (6)
- Other (specify) (7) __________________________________________________

Q5.4 Which of the following best describes the average number of medical emergencies the average PGY2 resident at your program responds to *by the end of their PGY2 year*?

- 1-25 (1)
- 26-50 (2)
- 51-99 (3)
- 100-149 (4)
- 150-199 (5)
- >200 (6)

Q5.5 By what ***time of the residency year*** is the average resident (July to June cycle) at your institution able to **independently *respond*** to medical emergencies?

- Beginning of residency-August (6)
- September-October (1)
- November-December (2)
- January-February (3)
- March-April (4)
- May-End of residency (5)
- Never able to respond independently (7)

| Page Break |
| --- |

Q5.6 Which of the following best describes how pharmacy residents are ***determined to be able to*** independently respond to medical emergencies at your institution? [**select all that apply**]

- Set time during residency year (e.g., Q3) (1)
- Meeting threshold of # of emergencies responded to (2)
- Passing grade on formal evaluation (e.g., simulation, exam) (3)
- Preceptor sign-off (4)
- Residents do not respond independently at my institution (5)
- Other (specify) (6) __________________________________________________

| Page Break |
| --- |

Q5.7 Please include any comments in this box detailing what resources were beneficial in developing a pharmacy resident medical emergency response system at your program.

________________________________________________________________

Skip To: End of Survey If Condition: Please include any comments... Is Displayed. Skip To: End of Survey.

End of Block: Demographics

Start of Block: No Response

Display This Question:

If Pharmacy residents in my PGY2 program actively participate (e.g., medication prep, clinical recom... = No

Q4.1 Which of the following describes why pharmacy residents do not actively participate in medical emergency response at your institution? [**select all that apply**]

- Does not fit into program's objectives (1)
- Lack of training (2)
- Pharmacists do not respond to medical emergencies at my institution (3)
- Other (specify) (4) __________________________________________________

Q4.2 Please include any comments in this box detailing what resources may be beneficial in developing a pharmacy resident medical emergency response system at your program.

________________________________________________________________

Skip To: End of Survey If Condition: Please include any comments... Is Displayed. Skip To: End of Survey.

End of Block: No Response

**Appendix B. Free-text Responses to Questionnaire**

**Q3.3 *Which of the following do you consider in identifying candidates (screening/ranking) for your PGY2 residency? [select all that apply] – Other***

CC

Participation in another emergency response team (e.g., stroke, sepsis, MERIT)

EM

Ability to apply experience from PGY1 emergency response to case discussion, also ability to discuss their role in the response team (with examples)

active vs. passive participation in medical emergencies

Certifications of PGY1 hospital (e.g. level 1/2 trauma, comprehensive stroke, etc to gauge acuity of patients typically cared for

hands-on code experience in any capacity

PGY1 EM rotation completed prior to applying

**Q5.1 *Which of the following describes the type(s) of medical emergencies that pharmacy residents respond to at your institution? [select all that apply] - Other***

Arctic Alert, STEMI Alert

Code stemi

Code violet

ED code 3 response, NICU response, OB RRT

massive transfusions, malignant hyperthermia

PERT

PRT, MTP, OB alert

residents respond in the same capacity as clinical pharmacists, no on-call

STEMI activations, procedural sedation in the ED

Toxicological Emergencies; Code STEMI; Acute Behavioral Emergencies

**Q5.3 *Which of the following training do PGY2 pharmacy residents complete at your institution to prepare them for code response? [select all that apply] – Other***

CC

3 part ACLS simulation program

ATLS

ENLS, PFCCS

EM

ATLS audit

ATLS, ENLS

In additional to ACLS/PALS we have department competencies

NRP

Weekly to monthly PALS mega codes

**Q5.6 *Which of the following best describes how pharmacy residents are determined to be able to independently respond to medical emergencies at your institution? – Other***

CC

ACLS certification

previously by preceptor sign off; starting this year with on call in house program, all residents (PGY1/PGY2) are responding independently from August forward once they are ACLS certified

No formal assessment

independent staffing shifts after training

preceptor sign-off for each individual rotation (ICU vs ED, etc)

A back up pharmacist is available in case meds from outside cart needed

ED pharmacists are available for support throughout duration of their coverage. Independent response is now (new this year) customized to each resident based on # emergencies attended, assessment of preceptor, and feedback of resident.

There is always another ICU/ED pharmacist on site as back up for an emergency response. They do not receive formal sign off versus a debrief and update after each scenario and resident update at our RAC mtg.

Complete checklist

PGY2s can respond independently usually by Nov-Dec. However, many times there is a preceptor at the code to check in with the PGY2 or to be an extra set of hands as a runner to Pyxis if the PGY2 is tied up in the room/with the code cart

Resident comfort

By Q2 they are main responder, but have a back up preceptor the entire year

EM

Completion of ER Competency Checklist

Generally, PGY1 residents do not respond independently outside of the ED rotation where they must have received preceptor approval by concesus of the preceptor group

Residents typically complete orientation activities and do a "buddy" call shift to shadow with another resident and after that point they are expected to be able to independently respond to emergencies

**Q5.7 *Please include any comments in this box detailing what resources were beneficial in developing a pharmacy resident medical emergency response system at your program.***

CC

ACLS course, Interdisciplinary Code Blue Simulation Trainings, Crash Cart Medication Training, Pharmacist Code Blue Pack Training

In situ simulation

Layered learning model: Q1 PGY-2 and CC/EM pharmacist respond with PGY-2 as learner, Q2-4 PGY-1 resident goes to code blue with PGY-2 CC/EM resident as main preceptor and CC/EM pharmacist as back up, all other responses Q2-4 are just PGY-2 CC/EM resident and C/EM pharmacist

Longitudinal topic discussion series, simulation lab

Most useful resource was feedback from our residents over the course of the years. Initial program developed based on preceptor experiences as residents.

We require med prep competency, crash cart review, minimum one code blue shadow, one active med prep, one documentation (code sheet), one pharmacy leader. Once all completed then may respond independently after RAC review and resident agreement

EM

developing a robust preceptor support system and training them on the code cart so they are familiar with the contents and can rely on muscle memory so they have the ability to think through the broader aspects fo the patient's condition.

None, pharmacist had already established this prior to starting the residency program. The residents follow the same model pharmacists had previously established.

pharmacist medication administration training and competency

pharmacist-led code training in a simluation center (with mannequin) that focuses on pharmacy-specific code response

Preceptors who have a ton of experience with this already

We recently implemented medical emergency simulation during PGY1 orientation (and PGY2 as appropriate/needed) to give the residents hands on experience in being the primary pharmacist at code blue and code stroke events
